# Boston criteria v2.0 for cerebral amyloid angiopathy without hemorrhage: An MRI-neuropathological validation study

**DOI:** 10.1101/2023.11.09.23298325

**Authors:** Aaron Switzer, Antreas Charidimou, Stuart J. McCarter, Prashanthi Vemuri, Aivi Nguyen, Scott A. Przybelski, Timothy G. Lesnick, Alejandro A. Rabinstein, Robert D. Brown, David S. Knopman, Ronald C. Petersen, Clifford R. Jack, R. Ross Reichard, Jonathan Graff-Radford

## Abstract

**BACKGROUND:** Updated criteria for the clinical-MRI diagnosis of cerebral amyloid angiopathy (CAA) have recently been proposed. However, their performance in individuals without intracerebral hemorrhage (ICH) or transient focal neurological episodes (TFNE) is unknown. We assessed the diagnostic performance of the Boston criteria version 2.0 for CAA diagnosis in a cohort of individuals presenting without symptomatic ICH.

**METHODS:** Fifty-four participants from the Mayo Clinic Study of Aging or Alzheimer’s Disease Research Center were included if they had an antemortem MRI with gradient-recall echo sequences and a brain autopsy with CAA evaluation. Performance of the Boston criteria v2.0 was compared to v1.5 using histopathologically verified CAA as the reference standard.

**RESULTS:** Median age at MRI was 75 years (IQR 65-80) with 28/54 participants having histopathologically verified CAA (i.e., moderate-to-severe CAA in at least 1 lobar region). The sensitivity and specificity of the Boston criteria v2.0 were 28.6% (95%CI: 13.2-48.7%) and 65.3% (95%CI: 44.3-82.8%) for probable CAA diagnosis (AUC 0.47) and 75.0% (55.1-89.3) and 38.5% (20.2-59.4) for any CAA diagnosis (possible + probable; AUC: 0.57), respectively. The v2.0 Boston criteria was not superior in performance compared to the prior v1.5 criteria for either CAA diagnostic category.

**CONCLUSIONS:** The Boston criteria v2.0 have low accuracy in patients who are asymptomatic or only have cognitive symptoms.. Additional biomarkers need to be explored to optimize CAA diagnosis in this population.

## Introduction

Cerebral amyloid angiopathy (CAA) is a common small vessel pathology, characterized by different degrees of cerebrovascular amyloid-β deposition in cortical and leptomeningeal vessels.^1^ CAA is a common cause of spontaneous lobar intracerebral hemorrhage (ICH), especially in the elderly and a key vascular contributor to cognitive impairment and dementia, including Alzheimer’s disease. The Boston criteria were developed as a set of clinical-MRI features to allow the diagnosis of CAA during life, in the absence of neuropathological analysis from post-mortem examination or biopsy (which still remains the gold standard for the definite diagnosis of the disease).^2^ Based on the presence of multiple strictly lobar cerebral microbleeds (CMBs), the initial iteration of the Boston criteria (v1.0), allowed for a diagnosis of probable CAA (i.e., indicative of probable underlying CAA pathology) primarily in patients with symptomatic ICH ^3^. The probable CAA diagnostic category remains the cornerstone for defining CAA in both the clinical and research setting.^2^ A modified version of the Boston Criteria (v1.5) further incorporated cortical superficial siderosis (cSS), a characteristic (and novel at the time) hemorrhagic marker of CAA, aiming to improve diagnostic accuracy.^4^

Advances in unraveling the much wider than previously appreciated clinical-MRI spectrum of CAA, coupled with important limitations of prior Boston criteria validation studies (i.e., small sample sizes, inclusion mainly of patients with prior ICH, and limited evaluation of the spectrum of CAA MRI markers^2,5^) prompted a large multicentre effort to update and validate the Boston criteria. The Boston criteria (v2.0) introduced non-hemorrhagic markers, white matter hyperintensity multi-spot (WMH-MS) pattern^6^ and MRI-visible perivascular spaces in the centrum semiovale (CSO-PVS),^7^ in addition to hemorrhagic markers, to improve sensitivity without significantly compromising specificity when compared to prior versions of the criteria.^8^

All previous versions of the Boston Criteria, including v2.0, were calibrated to identify more severe, bleeding-prone, cases of CAA. However, the performance of the Boston criteria v2.0 is uncertain in a population of cognitively unimpaired individuals and patients with MCI or dementia, presumably representing less severe CAA. There are few studies assessing the performance of any iteration of the Boston criteria in this population.^9,10^ Therefore, the aim of this study was to evaluate the diagnostic performance of the Boston criteria v2.0, in a population without symptomatic ICHs or transient focal neurological episodes (TFNE), which represented the majority of patients in the recent validation study.^8^

## Methods

### Study cohort and eligibility criteria

This is a retrospective cohort study including participants enrolled in either the Mayo Clinic Study of Aging (MCSA)^11^ or the Mayo Alzheimer’s Disease Research Center (ADRC). Participants were included if they had a MRI T2* gradient-recall echo sequence (GRE) and a brain autopsy with CAA evaluation. At the time of antemortem MRI, none of the participants had a history of ICH, TFNE, or CAA-related inflammation.

### MRI analysis

Participants had T1-weighted, T2 FLAIR, and T2*-GRE sequences on a 3-tesla General Electric MRI scanner between October 2011 and June 2016. Images were assessed for MRI features of CAA (cSS, WMH-MS pattern, CSO-PVS, and CMBs) according to international rating standards and recommendations^12,13^ and blinded to demographic, clinical and neuropathological data. Gradient recalled echo sequences were reviewed by a trained neurology fellow (ARS) and confirmed by a vascular neurologist (JGR).

Cortical superficial siderosis was defined as curvilinear hypointensities that track the cortical surface on T2*-weighted GRE, without corresponding hyperintensity on T1-weighted or FLAIR images.^14^ They were graded on the following scale: 0-absent, 1-focal (restricted to ≤3 sulci, or 3-disseminated (involving >4 sulci).^15^

The WMH-MS pattern was defined as >10 circular or ovoid WMHs surrounded by normal subcortical white matter on a FLAIR sequence.^6^ Images were reviewed by a trained neurology fellow (ARS), and each subcortical WMH spot was counted. If there were more than 10 WMH spots, the sign was considered present. WMH spots were not counted if associated with lacunar infarcts, perivascular spaces, or remote intracranial hemorrhages, confluent with a periventricular WMH, or confluent with another subcortical WMH. If the number of WMH spots was borderline (i.e., 10 or 11), the overall WMH pattern was used to determine whether the sign was present or not.

Perivascular spaces in the centrum semiovale were defined as round, oval, or linear shaped signal hyperintensity on T2-weighted sequences, or hypointensity on T1-weighted or T2 FLAIR sequences with intensity equal to cerebrospinal fluid.^16^ Most participants did not have T2-weighted fast spin echo (FSE) sequences; therefore CSO-PVS were counted on T1-weighted and T2 FLAIR sequences. CSO-PVS were counted following the Wardlaw method.^17^ Each axial slice was visually inspected and a representative slice with the most visible PVS was chosen in the centrum semiovale. The hemisphere with the most PVS was counted. If the PVS count was >20 in the centrum semiovale, the CSO-PVS sign was considered positive. Fifty-six percent of participants had both T2-FSE and T1-weighted sequences. In this subset of patients, CSO-PVS were counted on both sequences. T1-weighted sequences routinely underestimated the number of PVS compared to T2-FSE (**Supplemental Figure 1**). To maximize the number of participants in the study, PVS were counted on T1-weighted and T2 FLAIR sequences accepting that the true number may be underestimated.

Cerebral microbleeds were defined as a round hypointense lesion on T2*-weighted GRE that was distinct from iron or calcium deposition, or a vessel.^17^ CMB grading was performed by a trained imaging analyst and confirmed by a vascular neurologist (JGR). When it was unclear whether a lesion was a CMB or a vessel flow void, it was recorded as a possible CMB. Possible CMBs were not included in our analysis.^18^ An in-house, modified automated anatomic-labeling atlas was applied to the T1-weighted sequence and resampled into the GRE image on which the CMBs were graded.^19^ CMBs were classified as lobar only or deep/infratentorial.

A random sample of 20 participants were rated for WMH-MS pattern and MRI-visible severe CSO-PVS by ARS and SM. Interrater reliability showed excellent agreement (Cohen’s kappa 0.88 (95%CI: 0.64-1) for WMH-MS sign and 0.83 (95%CI: 0.61-1) for PVS). Participants were classified according to the Boston criteria v1.5^4^ and v2.0.^8^

Neuropathological grading

A board-certified neuropathologist (RRR) graded CAA severity based on Love Consensus Criteria.^20^ Each sample was formalin-fixed, embedded in paraffin, and stained using an amyloid-β antibody.^18^ CAA burden was rated in the parenchymal and leptomeningeal areas of each region on a scale of 0-3 (0=absent CAA; 1=scant CAA; 2=CAA in ≥2 arterioles with some circumferential Aβ; 3=widespread arteriolar CAA, many with circumferential Aβ). Capillary CAA was rated as present or absent. Forty-eight participants had complete CAA evaluation in all regions; the remainder had partial evaluations with missing data in the following regions: frontal *n*=3, temporal *n*=1, parietal *n*=2, occipital *n*=1, and hippocampus *n*=4.

### CAA classification and statistical analysis

Histopathologically verified CAA, used as the diagnostic reference standard for our analyses, was defined as at least one region with either parenchymal or leptomeningeal CAA graded 2 or higher according to the above scale. In secondary analyses, different CAA severity cut-offs were used to define the neuropathologic reference standard for CAA diagnosis, to explore if changes in the diagnostic performance of either set of criteria are different. Finally, in an explorative analysis of diagnostic performance, the presence of cognitive impairment, in addition to MRI markers, was required to apply the criteria for probable or possible CAA.

A participant’s baseline characteristics were summarized according to histopathologically verified CAA status using the mean and standard deviation for continuous variables and proportions for categorical variables. The diagnostic yield of the Boston criteria against the pathologically verified CAA reference standard was investigated using sensitivity, specificity, positive predictive value (PPV), negative predictive value (NPV), and area under the receiver operating characteristic (ROC) curve (AUC). Differences between the AUCs were tested using *Z*-tests from the R package pROC.^21^The level for statistical significance was set at 0.05 for all analyses. R version 4.3.1 was used for all analyses.

### Data Availability

The data that support the findings of this study are available from the corresponding author, upon request.

### Approvals

The Mayo Clinic Institutional Review Board approved this project. Patients provided written informed consent for the use of their medical information for research purposes.

### Reporting Guidelines

This study followed the STROBE reporting guidelines.^22^

## Results

Fifty-four participants (24% female, median age at MRI 75 years) were included in the study. Baseline characteristics are summarized in **Table 1**. The prevalence of moderate-to-severe CAA in at least one lobar region on brain pathology was 28/54 (52%). Forty-two participants had cognitive impairment (defined as a Mini-Mental Status Exam score <26). Based on the Boston criteria v2.0, 17 participants met criteria for probable CAA, and 10 met criteria for possible CAA diagnosis (**Table 1**). There was a high prevalence of lobar CMBs (33% of participants). Only 7% had either evidence of cSS or CSO-PVS. Sixty-six exhibited the WMH-MS pattern. Primary neuropathological diagnoses included Alzheimer’s disease, Lewy body disease, frontotemporal lobar degeneration, primary age-related tauopathy, argyrophilic grain disease, and prion disease.

**Table 1.** Demographic, clinical and imaging characteristics of study participants.

|  |  |
| --- | --- |
| Median age at MRI, years (IQR) | 75 (65-80) |
| Female, no. (%) | 13 (24) |
| Hypertension, no. (%)<sup>†</sup> | 25 (53) |
| Dyslipidemia, no. (%)<sup>†</sup> | 29 (62) |
| Diabetes mellitus, no. (%)<sup>†</sup> | 13 (28) |
| APOE ε4 carrier, no. (%) | 28 (53) |
| Education, years (SD) | 15.4 (2.8) |
| Mean MMSE at MRI, score (SD) | 19.3 (7.5) |
| Median age at death, years (IQR) | 77 (67-83) |
| Median time between MRI and autopsy, years (IQR) | 2.0 (1.4-2.7) |
| **Clinical diagnosis at MRI, no. (%)** |  |
| Cognitively unimpaired | 12 (22) |
| Mild cognitive impairment | 3 (6) |
| Dementia | 39 (72) |
| **Lobar CMB, no. (%)** |  |
| None | 36 (66) |
| 1 CMB | 8 (15) |
| 2 CMB | 6 (11) |
| 3 CMB | 1 (2) |
| 4+ CMB | 3 (6) |
| Deep CMB, no. (%) | 3 (6) |
| Cortical superficial siderosis presence, no. (%) | 4 (7) |
| WMH-MS present, no. (%) | 36 (66) |
| Enlarged CSO-PVS present, no. (%) | 4 (7) |
| **Primary neuropathological diagnosis, no. (%)** |  |
| Alzheimer's disease | 30 (56) |
| Lewy body disease | 11 (20) |
| FTLD-TDP | 7 (13) |
| FTLD-tau | 2 (4) |
| Primary age-related tauopathy | 2 (4) |
| Argyrophilic grain disease | 1 (2) |
| Prion disease | 1 (2) |

<sup>†</sup>Data missing from 7 participants.

|  | Histopathologically verified CAA |  |
| --- | --- | --- |
|  | Yes (n= 28) | No (n= 26) |
| **Boston criteria v1.5 diagnosis, no. (%)** |  |  |
| Probable CAA | 6 (21) | 5 (19) |
| Any CAA (Possible + Probable) | 8 (29) | 10 (38) |
| **Boston criteria v2.0 diagnosis, no. (%)** |  |  |
| Probable CAA | 8 (29) | 9 (35) |
| Any CAA (Possible + Probable) | 21 (75) | 16 (62) |

Histopathologically verified CAA refers to a grade of 2 or higher in at least 1 of the sampled regions based on the Love consensus criteria<sup>20</sup>. CAA=cerebral amyloid angiopathy; CMB=cerebral microbleed; WMH-MS=white matter hyperintensity multispot pattern; CSO-PVS=perivascular spaces in the centrum semiovale; FTLD=frontotemporal lobar degeneration; TDP=TAR DNA-binding protein 43; SD=standard deviation; IQR=interquartile range.

Diagnostic performance measures are summarized in **Table 2**. Using the Boston criteria v2.0, the sensitivity and specificity for probable CAA diagnosis (vs. non probable CAA) was 28.6% (95% CI: 13.2-48.7) and 65.3% (44.3-82.8) respectively, with an AUC of 0.47. Sensitivity and specificity for any CAA diagnosis (probable or possible CAA vs. no CAA) were 75.0% (95%CI: 55.1-89.3) and 38.5% (95%CI: 20.2-59.4) respectively, with an AUC of 0.57. There was no statistically significant difference in diagnostic performance between the Boston criteria v2.0 compared to v1.5 for probable CAA diagnosis (AUC 0.47 [95%CI: 0.34-0.60] vs. AUC 0.51 [0.40-0.62], *Z*= -0.94, *p*= 0.35) or any CAA diagnosis (AUC 0.57 [0.44-0.69] vs. AUC 0.45 [0.32-0.58], *Z*= 1.7, *p*= 0.09).

**Table 2.** Diagnostic performance of the Boston criteria v2.0 and v1.5 using histopathologically verified CAA as the reference standard.

|  | Probable CAA (vs. non-probable CAA) |  | Probable plus possible CAA (vs. no CAA) |  |
| --- | --- | --- | --- | --- |
|  | Boston criteria v2.0 | Boston criteria v1.5 | Boston criteria v2.0 | Boston criteria v1.5 |
| Sensitivity | 28.6% (13.2-48.7) | 21.4% (8.30-41.0) | 75.0% (55.1-89.3) | 28.6% (13.2-48.7) |
| Specificity | 65.3% (44.3-82.8) | 80.8% (60.7-93.5) | 38.5% (20.2-59.4) | 61.5% (40.6-79.8) |
| PPV | 47.1% (23.0-72.2) | 54.6% (23.4-83.3) | 56.8% (39.5-72.9) | 44.4% (21.5-69.2) |
| NPV | 46.0% (29.5-63.1) | 48.8% (33.3-64.5) | 58.8% (32.9-81.6) | 44.4% (27.9-61.9) |
| AUC | 0.47 (0.34-0.60) | 0.51 (0.40-0.62) | 0.57 (0.44-0.69) | 0.45 (0.32-0.58) |
| | $Z = -0.94, p = 0.35$ | | $Z = 1.7, p = 0.09$ | |

Comparison of paired ROC are reported as a Z-statistic and *p*-value. CAA=cerebral amyloid angiopathy; PPV=positive predictive value; NPV=negative predictive value; AUC=area under the receiver operator characteristic curve.

Using the Boston criteria v2.0 with probable CAA as the “rule in” diagnosis, changing the reference standard to histopathological grade of at least 1 (mild CAA), at least 3 (severe CAA), or presence of capillary CAA, did not significantly affect the diagnostic performance (**Table 3**). In a subset of participants with cognitive impairment (*n*= 42), the specificity of a “probable CAA” diagnosis improved at the expense of sensitivity (**Supplemental Table 2**).

**Table 3.** Diagnostic performance of the Boston criteria v2.0 for probable CAA vs. non probable CAA using different CAA neuropathologic grades of histopathologically verified CAA as the reference standard.

|  | Histopathological features |  |  |
| --- | --- | --- | --- |
| | ≥ Grade 1 CAA | ≥ Grade 3 CAA | Capillary CAA present |
| Sensitivity | 33.3% (19.6-49.6) | 30.0% (11.9-54.3) | 44.4% (21.5-69.2) |
| Specificity | 75.0% (42.8-94.5) | 67.6% (49.5-82.6) | 75.0% (57.8-87.9) |
| PPV | 82.4% (56.6-96.2) | 35.3% (14.2-61.7) | 47.1% (23.0-72.2) |
| NPV | 24.3% (11.8-41.2) | 62.2% (44.8-77.5) | 73.0% (55.9-86.2) |
| AUC | 0.54 (0.39-0.69) | 0.49 (0.36-0.62) | 0.60 (0.46-0.74) |
| Compared to<br>≥ grade 2 CAA | $Z = -0.73, p = 0.47$ | $Z = -0.20, p = 0.84$ | $Z = -1.3, p = 0.19$ |

Histopathological grades are based on the Love consensus criteria<sup>20</sup>. Comparison of paired ROC are reported as a Z-statistic and *p*-value. CAA=cerebral amyloid angiopathy; PPV=positive predictive value; NPV=negative predictive value; AUC=area under the receiver operator characteristic curve.

## Discussion

In this study, we explored the performance of the most recent iteration of the Boston criteria (v.2.0) in a population of individuals without symptomatic ICH or TFNE. Our main finding is that while the Boston criteria v2.0 improves sensitivity for CAA diagnosis (compared to v1.5), it comes with the expense of specificity to an almost similar degree. The net effect is that both the Boston criteria v2.0 and v1.5 show overall limited sensitivity and specificity and positive and negative predictive value in the setting of a population presenting without characteristic hemorrhagic CAA syndromes. The diagnostic accuracy and predictive value did not significantly improve when stratifying cases by varying grades of pathological CAA, nor did it change with the inclusion of cognitive impairment as a criterion for possible or probable CAA.

The performance in the population of our study is different from that observed in the recent validation study of the Boston criteria v2.0, which showed improved sensitivity (74.5%) and high specificity (95.0%) among patients who presented with spontaneous lobar hemorrhages, cognitive impairment, or TFNE (AUC 0.85).^8^ The Boston criteria were initially developed to determine the etiology of spontaneous ICH and to predict the risk of recurrent hemorrhage.^3^ Thus, the Boston criteria may be more accurate in and calibrated for identifying advanced (i.e., bleeding-prone) CAA pathology, which is more likely to cause symptomatic lobar ICH.^23^ Consistent with this hypothesis, a subgroup analysis in the Boston criteria v2.0 validation study showed low sensitivity (55.1%) and high specificity (96.2%) for predicting pathological CAA in cases presenting without ICH (but significantly improved compared to prior Boston criteria versions). Of note, patients in this subgroup analysis had TFNE or cognitive impairment and therefore differed from our study population as a whole.^8^ This is also reflected in the low prevalence of MRI markers of CAA in our population. Our data mirror a prior study that showed overall low sensitivity and specificity of the Boston criteria v1.0 in both hospital- and population-based settings of elderly individuals without ICH.^10^

The results of this study raise an important question about the clinical significance of incidental imaging biomarkers of CAA in asymptomatic people or patients presenting in a memory-clinic setting who have never experienced any symptomatic brain hemorrhage. Two large population-based studies showed a high rate of any pathological CAA on autopsy (78.9%) in participants without symptoms of CAA other than cognitive impairment.^9^ The Boston criteria v2.0 are very accurate in correctly identifying underlying CAA pathology in patients with symptomatic lobar hemorrhages or TFNE, but they may not accurately predict less severe pathological CAA changes in patients with minimal or no symptoms. Our findings require confirmation and further validation in larger population-based samples and patients with cognitive impairment, but they suggest that clinicians should exercise caution when applying the Boston criteria to an asymptomatic individual with a minimal incidental lobar CMB burden on brain MRI.

Our study has several limitations. A key limitation, inherent to all similar diagnostic validation studies in the field, is potential selection bias due to the requirement for neuropathological tissue. In combination with the relatively small sample size, this might reduce the generalizability of our findings. The use of GRE instead of SWI, which is less sensitive to blood-breakdown products and hence CMBs detection, may have increased the false negative rate in our analysis. There was a high degree of concomitant pathology in our population including Alzheimer’s disease, Lewy body disease, TDP-43, and 4-repeat tauopathy; these are associated with different prevalence rates and degrees of underlying CAA. Perivascular space rating was performed on T1-weighted sequences instead of T2-weighted sequences, which reduced our ability to detect CSO-PVS. Our study used the Love consensus criteria instead of the Vonsattel method used in the *Lancet* paper,^8^ which may limit the comparability of the two studies.

### Conclusion

The Boston criteria v2.0 are accurate when applied to patients with cardinal CAA hemorrhagic syndromes (symptomatic ICH or TNFE) and thus advanced disease, but the clinical relevance in asymptomatic individuals or patients with cognitive impairment and incidental radiological markers of CAA needs further exploration. Larger population-based studies should aim to assess our findings and determine the relationship between radiological markers of CAA and pathological severity. Additional biomarkers need to be identified to improve the detection of pathology on the milder end of the CAA spectrum.

